# Time, talk, and teamwork: Perceptions of personalised dementia care planning conversations in primary care

**DOI:** 10.64898/2026.02.20.26345977

**Authors:** Sarah Griffiths, Danielle Wyman, Mike Clark, Greta Rait, Nathan Davies

**Author notes:** **CORRESPONDING AUTHOR:** Sarah Griffiths.

## Abstract

**Background:** Dementia affects over 57 million people worldwide. UK and international policy position personalised, conversation-based care planning as central to post-diagnostic support. However, delivery in primary care is inconsistent, and many practitioners lack dementia-specific communication training. Existing evidence focuses on single roles or settings, leaving a gap in understanding how communication operates across the primary care workforce.

**Aims:** To identify what helps and hinders effective communication for integrated dementia care planning and determine the support and training needs of the wider primary care workforce.

**Methods:**

- Semi-structured interviews – 11 people with dementia, 13 family carers, and 19 primary care practitioners from diverse roles, exploring experiences of care planning conversations
- Reflexive thematic analysis

**Results:** Three themes were developed, progressing from micro-level communication practices (Theme 1: Beyond the tick-box), through triadic dynamics (Theme 2: Balancing voices in the conversation), to organisational influences (Theme 3: From silos to meaningful shared care planning). Time and Conversation as intervention cut across all themes, shaping trust and disclosure. Participants reported reliance on tick box approaches, inconsistent preparation, and uncertainty about care plan purpose and ownership. Non-clinical roles were commonly viewed as well placed to support meaningful conversations, but were often described as constrained by unclear remit and weak integration.

**Conclusions:** A persistent gap remains between policy ambitions and everyday practice. Time-pressured, checklist-driven encounters and fragmented systems undermine shared decision-making. The expanded primary care workforce offers untapped potential to address these gaps, but this requires clearer roles, formal integration, and targeted investment in communicative skills.

## BACKGROUND

Dementia affects around 57 million people worldwide, with nearly 10 million new cases each year [1]. As needs become increasingly complex, holistic care planning is essential for maintaining quality of life, autonomy, and dignity. The World Alzheimer’s Report 2022 [2] and NHS England [3] position personalised care and support planning as a core element of post-diagnostic dementia care, based on conversations about what matters most. People with dementia should have a personalised care and support plan, outlining their needs and how they will be addressed, reviewed annually and shared across services [3].

Key principles of personalised care planning include prioritising conversation over form-filling, ensuring adequate preparation, and supporting partnership and shared decision-making [4]. Recent UK policy reinforces this emphasis, calling for co-produced, holistic care plans alongside a shift to integrated, community-based services and shared digital access to care plans [5].

Despite these ambitions, routine dementia care planning in primary care is often inconsistent. The Quality and Outcomes Framework (QOF) annual dementia review [6] is a potential general practice mechanism for personalised care planning, yet evidence indicates wide variation in the quality and delivery [7, 8]. Policy therefore encourages wider team involvement, including roles such as social prescribers, care coordinators, and dementia advisors [3]. While these roles provide personalised, strengths-based support, they often operate separately from QOF reviews, limiting continuity and shared ownership. In England, the Additional Roles Reimbursement Scheme (ARRS), funded by NHS England, has expanded the primary care workforce by more than 26,000 staff [9], many of which could support dementia care planning. However, implementation is uneven, with reported gaps in preparation, retention, and team integration [9].

Communication in dementia care is complicated by cognitive, linguistic, and motor speech changes [10], and many practitioners report low confidence when communicating with people with dementia [11]. While general personalised care training exists [12,13], there is little dementia-specific guidance, creating a gap between policy expectations and available support.

Existing research offers useful insights into the communication experiences and training needs of practitioners delivering personalised dementia care, but the evidence base remains limited. A recent review [14] found little evidence on communication strategies for personalised dementia care in primary care, with most studies focusing on advanced dementia in care homes. Studies of social prescribers show reliance on carers for communication and a tendency to consult carers before engaging directly with people with dementia [15], while research on dementia support workers highlights challenges in delivering remote personalised care, including difficulties using visual communication tools [16]. Although valuable, these studies focus on single practitioner groups or specific delivery modes, and do not capture the range of communication experiences across the expanding primary care dementia workforce or routine face-to-face care planning. This gap is significant given that personalised care planning depends on practitioners’ ability to engage people with dementia and carers in meaningful, values-based conversations. As dementia care is increasingly distributed across teams, a broader, multi-perspective understanding of what supports or hinders effective communication is needed. This study therefore examines personalised dementia care planning conversations from the perspectives of people with dementia, carers, and diverse primary care practitioners to inform workforce development.

## AIMS

This study aimed to: (1) identify what helps and hinders effective communication for integrated dementia care planning, and (2) determine the support and training needs of the wider primary care workforce to facilitate high-quality personalised dementia care planning conversations.

## METHODS

We conducted a qualitative interview study using reflexive thematic analysis [17]. Qualitative methods are reported in line with COREQ [18].

### Ethics

The study received Health Research Authority approval and ethical approval from Wales REC 4 (24/WA/0167) on 17 June 2024. Capacity to consent among participants with dementia was determined by an experienced researcher (SG) in line with the Mental Capacity Act [19]. Written or verbal informed consent was obtained from all participants. In line with the approved procedure, where verbal consent was obtained, this was audio recorded, witnessed and documented in writing by a researcher. The document and audio file were stored on a secure university data management system.

### Public Involvement

This study was co-developed with a lived experience group [20]. Two people with dementia, one carer, and one former carer collaborated with the team throughout, advising on ethics, recruitment, methods, and materials, and contributing to data interpretation, guided by research on involving people with dementia in analysis [21].

### Recruitment

Recruitment took place between 22^nd^ August 2024 and 3^rd^ April 2025. We aimed to recruit up to 12 people with dementia, 12 carers, and 26 primary care practitioners across England. Practitioners were eligible if they worked in or with primary care and were involved in personalised dementia care planning. The sample size was chosen to ensure sufficient ‘information power’ [22] across groups to address the research questions. Eligibility criteria are shown in Table 1.

**Table 1:** Eligibility Criteria.

| Participant Group | Inclusion Criteria | Exclusion Criteria |
| --- | --- | --- |
| People with Dementia | • Clinical diagnosis of dementia, confirmed by practitioner at study site<br>• Aged ≥18<br>• Mild–moderate dementia with capacity to consent<br>• Able to communicate in English<br>• Has engaged with a primary care practitioner for a care and support planning conversation* within the past month (timeframe selected to counteract recall difficulties) | • Under 18<br>• Lacking capacity to consent<br>• Unable to communicate in English |
| Carers | • Adult family member or friend providing unpaid care to a participating person with dementia<br>• Able to communicate in English<br>• Willing and able to provide written or verbal informed consent<br>• Has engaged with a primary care practitioner for care and support planning with the person with dementia within the past month | • Under 18<br>• Carers with a cognitive impairment<br>• Paid carers not related to the person with dementia |
| Primary Care Practitioners | • Practitioners involved in personalised dementia care planning in primary care* (e.g., GP, practice nurse, dementia advisor, social prescriber, specialist dementia nurse, care coordinator)<br>• Able to communicate fluently in English | • Under 18 |
|  | • Willing and able to provide written or verbal informed consent |  |

\*This was explained to potential participants as any conversations designed to gain an understanding of the support needs and strengths of a person with dementia +/- their carers.

Recruitment sources included Primary Care Networks (groups of general practices sharing resources to deliver integrated care), charities supporting people with dementia, professional networks and social enterprises employing practitioners such as social prescribers and dementia advisors.

### Data collection

Semi-structured interviews were conducted in person or via video call, and individually or as dyads, depending on participant preference and practical considerations. Topic guides (Supplementary File) were developed by the research team and informed by literature [14, 23], policy [3] and input from the lived experience group. Interviews with people with dementia and carers explored their experiences of personalised care planning conversations, including what mattered to them, helpful or unhelpful communication approaches, preferences, and perceived gaps. Practitioner interviews focused on experiences of facilitating these conversations, including challenges, strategies, cross-service integration, and training needs.

### Data analysis

Interviews were audio-recorded, transcribed verbatim, and analysed using reflexive thematic analysis, led by SG. Following Braun and Clarke’s guidance [17], analysis moved iteratively through familiarisation, inductive coding, and the development, review, and refinement of themes, with repeated transcript readings to ensure deep engagement. Initial codes were generated systematically across the dataset.

The project adopted an experiential, constructionist stance, treating participants’ accounts as meaningful while acknowledging their contextual shaping. Coding attended to both semantic and latent meanings. To enhance rigour, DW independently reviewed a subset of transcripts and coding to support interpretation and reflexive discussion [24]. Coding and theme development were refined through regular team discussions with all authors and the lived experience group, whose reflections informed analytic decisions and strengthened the coherence and credibility of the final themes.

## RESULTS

### Participants

Forty-three semi-structured interviews were conducted with 11 people with dementia and 13 family carers with varied diagnoses and living arrangements, and 19 practitioners from a range of roles. Participants took part individually, as dyads, or in one triad (Tables 2–3). The practitioner sample mostly had under five years’ experience, reflecting the relative newness of many primary care roles (Table 3).

**Table 2:** Participant characteristics – people with dementia and carers.

| Participant | ID | Region | Sex | Age | Ethnicity | Type of dementia<br>(Interviewee or<br>cared for<br>person) | IMD<br>quintile | Individual<br>interview/dyad |
| --- | --- | --- | --- | --- | --- | --- | --- | --- |
| Person with dementia | DE-01 | SE England | F | 76-80 | White Other | FTD language variant - Primary progressive aphasia | 5 | Dyad |
| Carer | CE-01 | SE England | M | 71-75 | White British |  | Same as PWD |  |
| Person with dementia | DE-02 | North England | M | 61-65 | White British | FTD language variant - Primary progressive aphasia | 5 | Dyad |
| Carer | CE-02 | North England | F | 61-65 | White British |  | Same as PWD |  |
| Person with dementia | DE-03 | North England | M | 71-75 | White Irish | Vascular | 1 | Individual interview |
| Person with dementia | DE-04 | North England | M | 71-75 | White British | Alzheimer's | 1 | Individual interview |
| Person with dementia | DL-03 | SE England | M | 81-85 | White British | Alzheimer's | 4 | Dyad |
| Carer | CL-03 | SE England | M | 56-60 | White British |  | 5 |  |
| Person with dementia | DL-06 | SE England | M | 76-80 | White British | Unspecified | 4 | Dyad |
| Carer | CL-06 | SE England | F | 76-80 | White British |  | 4 |  |
| Person with dementia | DL-01 | SE England | F | 76-80 | White Irish | Unspecified | 5 | Individual interview |
| **Person with dementia** | DL-04 | SE England | M | 86-90 | White Other | Mixed | 3 | Triad |
| **Carer** | CL-04 | SE England | F | 86-90 | White Other |  | Same as PWD |  |
| **Carer** | CL-05 | SE England | F | 56-60 | White Other |  | 3 |  |
| **Person with dementia** | DE-06 | South England | M | 61-65 | White British | Lewy Body | 4 | Individual interview |
| **Person with dementia** | DD-06 | SW England | M | 91-95 | White British | Vascular | 4 | Dyad |
| **Carer** | CD-06 | SW England | F | 86-90 | White British |  | Same as PWD |  |
| **Person with dementia** | DE-05 | North England | F | 46-50 | British Asian – Pakistani | Alzheimer's | 1 | Individual interview |
| **Carer** | CL-01 | SE England | M | 36-40 | British Asian – Bangladeshi | Unspecified | 3 | Individual interview |
| **Carer** | CD-01 | SW England | M | 71 - 75 | White British | Unspecified | 2 | Individual interview |
| **Carer** | CD-02 | SW England | F | 76-80 | White British | Fronto-temporal behavioural variant | 3 | Individual interview |
| **Carer** | CD-03 | SW England | F | 81-85 | White British | Mixed | 2 | Individual interview |
| **Carer** | CL-02 | SE England | F | 56-60 | White Irish | Mixed | 3 | Individual interview |
| **Carer** | CL-07 | SE England | F | 51-55 | White British | Alzheimer's | 1 | Individual interview |

**Table 3:** Participant characteristics – Practitioners.

| Role | ID | Region | Sex | Ethnicity | Work setting | Years experience | Ethnicity |
| --- | --- | --- | --- | --- | --- | --- | --- |
| **Social Prescriber** | PE-01 | Southeast England | F | White Irish | GP surgeries | <5 | White Irish |
| **Dementia Advisor** | PL-01 | Southeast England | M | Black British | Charity | 11-20 | Black British |
| **Social Prescriber** | PE-02 | Southwest England | F | White British | GP surgeries | <5 | White British |
| **Social Prescriber** | PE-03 | North of England | F | White British | GP surgeries | <5 | White British |
| **Social Prescriber** | PD-01 | Southwest England | F | White British | Social Enterprise working with GP surgeries | <5 | White British |
| **Social Prescriber** | PD-02 | Southwest England | F | White British | Social Enterprise working with GP surgeries | 5-10 | White British |
| **GP** | PL-02 | Southeast England | F | White British | GP surgery | 5-10 | White British |
| **GP** | PL-03 | Southeast England | M | Mixed Caribbean | GP surgery | <5 | Mixed Caribbean |
| **GP** | PL-04 | Southeast England | F | White other | GP surgery | <5 | White other |
| **Dementia Advisor** | PL-05 | Southeast England | F | White British | Charity | <5 | White British |
| **Admiral Nurse** | PE-04 | North of England | F | White British | GP surgeries | <5 | White British |
| **Dementia Advisor** | PL-06 | Southeast England | F | White British | Charity | <5 | White British |
| **Care co-ordinator** | PL-07 | Southeast England | F | White British | GP surgery | 5-10 | White British |
| **Admiral Nurse** | PE-05 | Southeast England | F | White Other | GP surgeries | <5 | White Other |
| **GP** | PL-09 | Southeast England | F | White British | GP surgery | >20 | White British |
| **Dementia Advisor** | PL-08 | Southeast England | F | White British | Charity | <5 | White British |
| **GP** | PL-10 | Southeast England | F | White British | GP surgery |  | White British |
| **Trainee GP**<br>**and**<br>**Clinical**<br>**Academic**<br>**Fellow** | PD-05 | Southwest<br>England | F | White British | GP surgery | <5 | White British |
| **Dementia**<br>**Advisor** | PE-07 | North of<br>England | F | British<br>Pakistani | Charity | <5 | British<br>Pakistani |

### Findings

Three core themes and eight sub-themes were developed (Fig. 1), broadening in scope from micro-level communication practices (Theme 1: Beyond the tick-box), through the dynamics of balancing voices (Theme 2: Balancing voices in the conversation), to wider contextual influences on care planning (Theme 3: From silos to meaningful shared care planning). ‘Time’ and ‘Conversation as intervention’ were identified as cross-cutting themes and are integrated across the findings.

**Figure 1:**
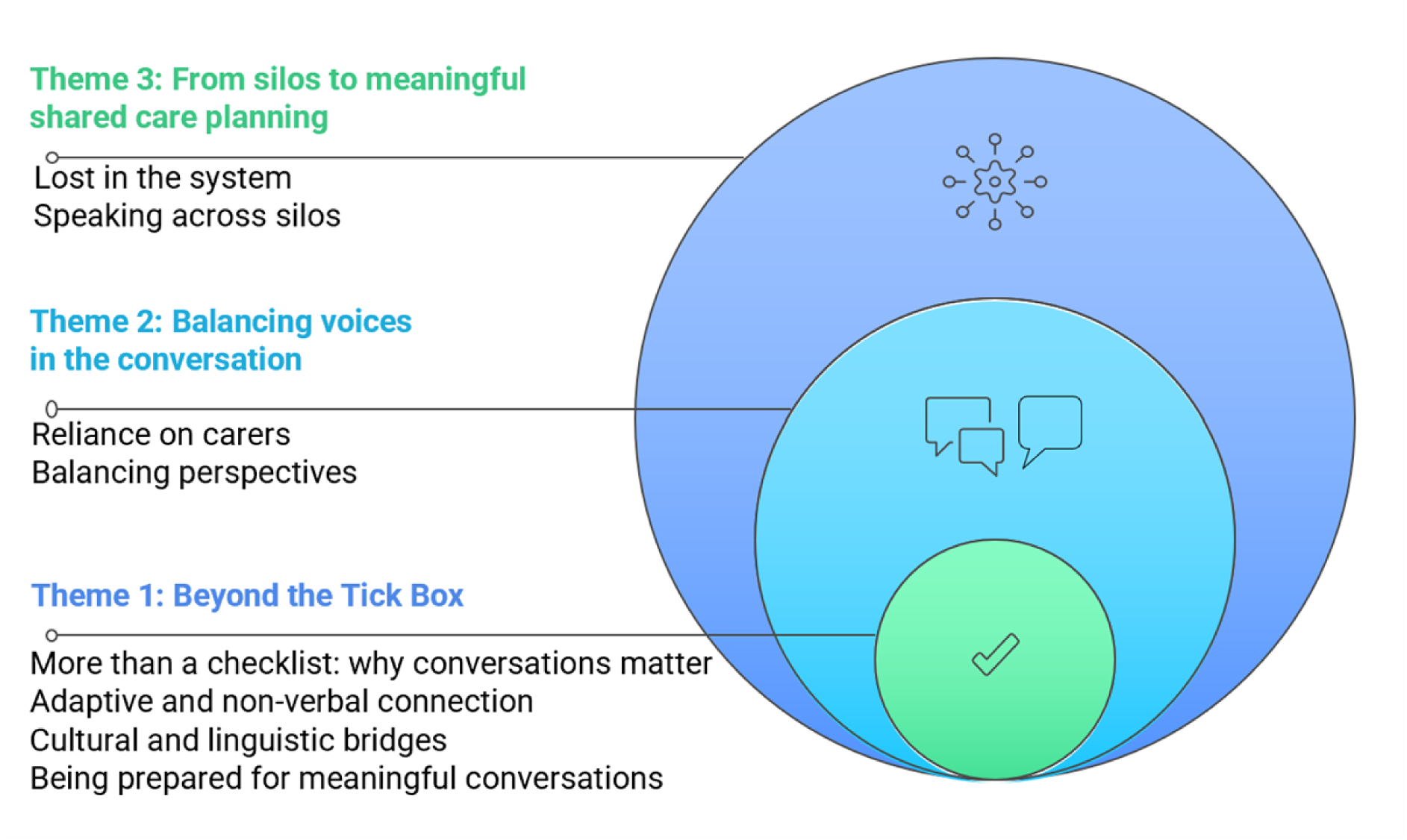
Themes.

### Theme 1: Beyond the tick box

This theme examines how checklist-driven approaches often limit connection and personalisation. Participants felt meaningful conversations happened when both sides had time and preparation, communication adaptations were adopted to support communication (e.g., visual resources), deepening understanding and supporting genuine partnerships.

#### More than a checklist: why conversations matter

Most practitioners relied on standard checklists, which practitioners, people with dementia and carers recognised as impersonal and limited in capturing the complexity and fluctuation of dementia.

> *‘It was an entire tick box exercise whereby it was merely yes or no, and even if it was a no, it wouldn’t have made any difference, there was no follow-on from the no.’* (PWD: DE-06)
>
> *‘If you go in with too much structure some people just won’t engage…you won’t get the information you need. So, you’ve got to do it in a…communicative way.’* (Social prescriber: PE-02)

Participants across all groups felt that meaningful conversations required time, a relaxed pace, and genuine curiosity, with unhurried encounters helping them feel known, creating space for honest reflection, and enabling deeper discussion through open-ended questions.

> *‘Questions like, “And you’re coping OK?”. They’re dead ends. Better to ask, “What’s your day like?” or “When did you last do something for yourself?’”* (Carer:CL-02)

They also noted that setting shaped interaction, with home visits offering fuller context, clinics fewer distractions, and remote consultations often feeling rushed. Flexibility was valued over any one format.

> *‘When I have done home visits the person is much more relaxed…they’re able to talk about themselves more. On the telephone sometimes people with dementia just clam up.’* (Dementia Advisor:PL-06)

Time enabled people to share their stories, helping them process change, build trust, and enabling practitioners to better identify strengths and support needs. When asked what practitioners should ask to understand his needs and strengths, one person with dementia said:

> *‘My life history –where I was born, who I married, and what happened to them.’* (PWD:DL-03)

Reassurance was a valued outcome of conversation, with affirmation and encouragement reducing anxiety and guilt for both people with dementia and carers.

> *‘I worry and feel guilty about doing the right thing…She said buy him what he’ll eat at this stage. It was totally reassuring and so lovely.’ (Carer:CL-01)*

People with dementia and carers felt checklists often missed important areas like their everyday lives, how well they understand their medications, or planning for contingencies, reinforcing a sense of being unseen.

> *‘I get asked, have you had any falls? But they could ask – Am I still able to go shopping? …Am I still able to use the computer?’* (PWD:DE-04)

Moving away from checklists towards a conversational approach enabled collaborative problem-solving, with gentle, scenario-based examples helping to normalise support and make change feel manageable. Speaking about a social prescriber, one person with dementia reflected:

> *‘He didn’t just say, “Get a carer.” He said, “How would you feel about someone coming in with a cup of tea, to check you’re alright?” It made it easier to accept.’* (PWD:DE-05)

#### Adaptive and non-verbal connection

Participants across all groups emphasised that non-verbal cues shaped whether people felt safe and understood, with small gestures and a calm presence helping to build trust.

> *‘She’s very friendly… she’ll just sort of like…touches you… she really looks at you.’* (Carer:CE-02)

For some people with dementia, emotional tone mattered more than the specific content of what was said, serving as an immediate indicator of whether an interaction felt safe or threatening.

> *‘When I’m speaking to somebody, straight away I’m trying to analyse: are they a threat?’* (PWD:DE-03)

Some practitioners described adapting their communication by simplifying language and using visual or creative strategies to make abstract information more concrete, support comprehension, and provide reassurance:

> *‘If I’m talking about something new or abstract… I print off details… if there’s photographs of people [in a support group], that might give reassurance.’* (Social Prescriber:PD-01)

Others described wanting more training in these approaches, alongside clearer permission and remit to use them in practice.

> *‘Training on… pictorial stuff…would [be] really helpful…knowing what we can use.’* (GP:PL-02)
>
> *‘A creative [resource] with…a map of their life to show also they’re still significant… I’d love to do it but from a commissioners standpoint, they [wouldn’t] think that’s your remit.* (Social prescriber:PD-01)

#### Cultural and linguistic bridges

Culture and language shaped whether conversations felt safe and inclusive, with participants valuing practitioners who showed cultural humility and respect.

> *‘If you were to give them respect by saying ‘Salam Alaikum’ because you knew they were Pakistani, you would see that smile.’* (Dementia Advisor: PE-07)

Some practitioners avoided potentially sensitive subjects, such as sexuality or gender, for fear of causing offence.

> *‘…we exclude questions…about being gay, because these kinds of things are forbidden in Islam.’* (Dementia Advisor:PE-07)

While well-intentioned, this could lead to important parts of identity being overlooked, suggesting the need for respectful and more conversational ways of exploring identity.

Language barriers were a further challenge. Telephone interpreters were often inaccessible, face-to-face bilingual support inconsistent, and practitioners relied on creative strategies such as involving relatives to sustain conversation or using drawing and pictures.

> *‘One lovely guy …was telling me about when he was young…I caught the word and looked it up, showed him pictures. It was beautiful…like a big castle.’* (Social Prescriber: PE-01)

Practitioners who shared cultural lived experiences with people with dementia were also seen as powerful bridges, reducing dementia stigma within communities.

> *Being prepared for meaningful conversations*

Preparation was seen as essential for conversation that feels respectful, supportive, and genuinely personalised. People with dementia and carers valued knowing what to expect and having time to reflect before reviews, helping them identify priorities and participate actively rather than reactively.

Practitioners emphasised the importance of preparing for the encounter, by setting expectations: introducing their role and the purpose of the conversation to avoid confusion and build trust.

> *‘I explain to them I’m not a nurse or a doctor… I just say, I’ve come to see how you are.’* (Dementia Advisor:PD-07)

Knowledge of dementia subtypes and trajectories was also viewed as part of being adequately prepared for personalised care planning, with gaps in understanding undermining trust.

> **Carer:** ‘They all think he’s got Alzheimer’s cos dementia is just Alzheimer’s’ **PWD:** ‘Yeah, that’s true.’ **Carer**: *‘It’s PPA and they don’t understand that. They should do but they don’t’* (Dyad: DE-02; CE02)

Overall, this theme underscores the importance of time and the idea that conversation itself is an intervention and a foundation for personalised care. Participants valued care-planning not as a procedural task, but as a flexible, relational process through which reassurance, shared understanding, and personalised support are built.

### Theme 2: Balancing voices in the conversation

Central to this theme is the challenge of ensuring both people with dementia and carers feel heard during care-planning conversations.

#### Reliance on carers

Time pressures and inaccessible formats (such as phone calls) often led practitioners to defer to carers, sidelining people with dementia. Carers felt pressured to speak on their behalf and worried about misrepresenting their views.

> *‘…on the phone…you’re having to be really careful cos it’s my interpretation of what’s going on…it’s not necessarily correct, is it?’* (Carer:CE-02)

Some practitioners described mistrusting carer input in sensitive areas, adding complexity to triadic care planning, as concerns (e.g., about driving or behaviour) were sometimes minimised to avoid upsetting the person with dementia or triggering consequences such as sedatives or care transfers.

Theme one explored how language barriers shape conversations. When family members support communication, practitioners questioned whose voice was being heard when relatives declined professional interpretation without consulting the patient:

> *‘Is everything you’re saying via the family member being directly translated or is it their take?… Sometimes we say, “Would you like me to get a telephone translator?” but instead of that being translated to the patient, the answer is just no.’* (GP:PL-01)

Language barriers not only complicate communication but can also contribute to the exclusion of person with dementia from care planning conversations, amplifying issues of voice and representation in care.

#### Balancing perspectives

Carers often felt rushed or conflicted when their views differed from the person with dementia’s or when appointments left little space for their own needs. Both parties could struggle with honesty in each other’s presence, highlighting the need for sensitive management, including awareness of family dynamics, separate conversations, and skills to balance perspectives.

> *‘I say, “Oh, is that definitely what’s happening at the moment…it might have been like in the past, but what about now, and…invite more of a group discussion… there are times where…it’s easier to speak to them separately.’ (GP: PL-02)*

Carers also wanted recognition that their cared for person’s account may not fully capture the situation, without feeling they must work hard to justify their perspective.

> *‘it’s being able to have two conversations… some [practitioners] are skilful at that. You don’t come away thinking “Oh, God, my mother said X, Y, and Z, and that’s totally not the case.” You don’t know if they’ve… taken that…as gospel.’* (Carer:CL-02)

Some practitioners encouraged families to discuss issues privately first, supporting preparation for care planning (Theme one) and enabling later separate conversations. They emphasised the importance of careful question framing in triadic interactions, to protect dignity and balance the needs of all parties.

> *‘When a [practitioner] is directing questions at the carer in front of the person with dementia, they should be sensitive… especially when…delicate, regarding incontinence. They could frame [as] “What are your worries for your loved one regarding this area?”* (Carer:CL-02)

Practitioner accounts of complex situations highlight the skilled work required for listening to and acknowledging both perspectives:

> *‘Maybe 10 years ago they said don’t ever let me go into a nursing home…they’re now realising they can’t cope…listening to them both and advocating for what’s safe and best for the patient can be difficult…Listen to what people are saying and try to guide them in whichever direction you feel they want to go, and you think is best for them.’* (GP:PD-05)

However, some practitioners described facing challenges when carers decline support despite signs of strain or risk, and when carers struggled to accept changes in the person with dementia or interpreted symptoms as deliberate behaviour.

> *‘Sometimes carers… think [PWD is] doing something on purpose… They might not recognise there’s been a change… and this is the reason for it.’* (Dementia Advisor:PL-08)

Including carers as equal partners in care planning therefore involves recognising their level of understanding, acceptance and emotional strain.

Effective care planning depends on taking time to negotiate how both the person with dementia and the carer are heard. Conversation itself can be experienced as an intervention that validates and supports each perspective.

### Theme 3: From silos to meaningful shared care planning

This theme shows how aspirations for personalised dementia care are undermined by persistent fragmentation. *‘Lost in the system’* concerns how this fragmentation is felt, through communication gaps that leave people overwhelmed and uncertain about care planning and who should be involved. ‘*Speaking across silos’* examines the communicative mechanisms through which fragmentation is sustained and shows how communication that bridges organisational boundaries is fundamental to meaningful, person-centred care planning.

#### Lost in the system

Siloed working and disjointed systems were described as the norm across the sample. GPs felt overwhelmed by workload and time pressures, limiting collaboration despite recognising its value, while non-clinical practitioners reported feeling disconnected and unclear about roles and how to work together.

> *‘Understanding how all the different services work together would have been useful… it needs to be much more cohesive and joined up for it to be really helpful.’* (Dementia Advisor:PL-06)

The absence of a named point of contact for patients/carers compounded this sense of disconnection. Non-clinical team members often assumed coordination roles by default, trying to bridge systemic gaps and provide reassurance, but without formal recognition or support.

> *‘I’m finding…the carers in particular struggling so much they’re overwhelmed with contacts. So, I’m the one person who’ll be the link to all the other contacts, they will call me first.’* (Social Prescriber:PE-01)

For some, this carried an emotional toll, requiring repeated advocacy within an unresponsive system.

> *‘It’s a frustration. You just have to keep poking the bear until somebody actually listens …sometimes it may be too late…the patient may have had a fall…it’s really a stopping of…an ultimate horrific end.’* (Care Co-ordinator: PL-07)

For people with dementia and carers, the consequences of fragmentation were evident in everyday encounters. They described interacting with multiple practitioners without knowing who they were or how they were connected, creating a sense of being processed rather than held within a coherent care network.

> *‘**Carer:** We had this guy phone from…What did they call him? He was from the clinic…but he wasn’t a doc-…Not a therapist, even… He phoned twice and had a chat with you and then discharged you. **Person with dementia:** It didn’t do anything. It was just filling up the NHS “Oh, now this is somebody else that you’ll be speaking to.”’* (Dyad:DL-06 and CL-06)

There was a general conceptual confusion over care plans across all participants. Many people with dementia and carers did not recall ever having seen a care plan or participated in a dementia review. Care plans were rarely referred to or explained in accessible terms.

> *‘I wonder if it was that lady that came here…she might have been doing the care plan…I think they need to be clearer; written down.’* (Carer: CL-04)

Some practitioners were also unclear about the concept of a care plan and whether what they were doing constituted care planning.

> ‘*I’ve never heard of anyone talk about that before to be honest*.’ (PD-02: Social Prescriber)

People with dementia called for more collaborative, two-way models that recognise their ongoing agency; models that, in principle, should already be in place.

> *‘Monitoring should be a two-way engagement…knowing that if we’ve had issues we would go back and ask for a meeting together and then it would grow from there.’* (PWD:DE-06)

These accounts show that people with dementia, carers and practitioners are not just navigating a fragmented system but often feel lost within it, with communication gaps and unclear roles and processes leaving them overwhelmed, frustrated, and unsupported.

#### Speaking across silos

Across accounts, Collaboration within primary care and with other services was constrained by incompatible systems and limited time and space for multidisciplinary dialogue. People with dementia noticed when practitioners could access their records, avoiding repetitive or impersonal assessments and improving interaction quality.

> *‘It went well…[they] knew about me so didn’t have to go through…repetitive statements which means…they haven’t really read the medical record beforehand.’* (PWD:DE-06)

However, even when practitioners shared access to electronic care records, they lamented the lack of conversations with each other, describing fragmented, task-oriented exchanges that limited shared sense-making.

> *‘It doesn’t feel joined up in terms of a multidisciplinary team…like an urgent email referral into older people’s mental health team…they look at my notes, and I see their notes…but there would be no direct communication…we don’t have time to review [electronic] feedback from every service we refer to… that…loop of communication just physically can’t happen.’* (GP:PD-05)

As a result, care plans were rarely dynamic or genuinely shared, becoming annual, template-driven reconstructions disconnected from evolving needs.

Many practitioners described teams working in parallel, with heavy caseloads and little visibility of each other’s work, making it hard to sustain a shared sense of direction.

> *‘… other teams are just as busy…I haven’t always got time to check what they’re doing… if we had clear care plans that would be so much better because we’d all know…the direction of travel.’* (Admiral Nurse:PE-05)

Referrals between services were inconsistent and often lacked essential details, placing the burden of discovery on those picking up new cases.

> *‘We often get limited information…It just says can you speak to this person with dementia…You pick up the telephone…that’s when you start to realise what you’re dealing with.’* (Dementia Advisor:PL-06)

Formal structures for shared planning were limited, and information-sharing often depended on ad hoc communication.

> *‘It’s more corridor conversations… “Oh, I’ve seen so-and-so, they might be struggling a bit with that.”’* (Admiral Nurse:PE-05)

Structural silos also shaped how the capacities of non-clinical roles were used or overlooked. Many described being invisible within the system, with GPs unaware of their existence even when efforts had been made to introduce their roles.

> *‘Half [the GPs]… don’t even know we exist…even though I left leaflets in the surgery.’* (Dementia Advisor:PE-07)

Local variation meant some people with dementia and carers formed strong relationships with non-clinical practitioners, while others never encountered them. Even within multi-disciplinary teams (MDTs), these roles were often treated as peripheral, despite their capacity to build trust and support values-based conversations. Practitioners expressed a desire for greater connection, clearer expectations, and more meaningful involvement.

> *‘Social prescribers have time… I’d like us to be more connected to clinicians…Our job is to make their job better… We sit and talk…most doctors would love that luxury. If they guide us on what they want… what do you want us to do? What are you trying to achieve here …getting it nailed what this care plan is supposed to achieve.’* (Social Prescriber: PE-01)

Despite this potential, non-clinical roles often lacked access to records or formal channels to feed their insights into care planning.

Where co-location, shared technology, or regular MDT meetings existed, practitioners reported more fluid collaboration, clearer communication, and greater confidence in shared decisions.

> *‘The GP might ask us to cover a particular topic with somebody…we can go to the house…sit with them…help them think through scenarios.’* (Social Prescriber:PE-03)

Together, these accounts show that meaningful shared care planning depends on both structural coordination and time and space for practitioners to talk, reflect, and plan together. What was often missing was sustained conversation, between teams and with people with dementia and carers, capable of turning fragmented activity into coherent, person-centred care. Where enabled, such dialogue can bring clearer direction, greater confidence, and reduced emotional strain, positioning conversation itself as a therapeutic and integrative intervention.

## DISCUSSION

Findings illuminate what helps and hinders effective communication for integrated dementia care planning and identify the support and training needs of the wider primary care workforce. There is a persistent gap between policy expectations for personalised care and everyday practice. Although guidelines prioritise conversation over form-filling [4], care planning was often experienced as checklist-driven, constraining meaningful dialogue. Given people with dementia may conceal or downplay difficulties to avoid stigma and preserve dignity [25], such formats risk obscuring what matters most.

Time operated as a critical enabler of effective communication, allowing unhurried discussions in which participants could share stories, reflect on change, and build trust. Conversation itself was experienced as a form of intervention, offering reassurance, partnership, and emotional support rather than merely gathering information. These time-rich interactions were often linked to non-clinical roles perceived as having greater relational capacity, yet their informal and marginalised status limited their impact on integrated care planning.

Despite being central to personalised care [4], preparation for care planning conversations was often inadequate, leaving their purpose, scope, and roles unclear and undermining trust. Participants stressed that meaningful involvement of people with dementia requires adapted communication to support memory and comprehension, yet many lacked the time, training, or tools to provide this. Wider workforce roles were often seen as better positioned to offer such support, but without formal endorsement or training, their potential remains underdeveloped.

Participants also noted cultural and linguistic dimensions of care planning. Small, culturally attuned gestures were seen to support rapport, while language barriers and avoidance of sensitive identity-related topics risked excluding what mattered most.

Practitioners valued a conversational, collaborative approach to care planning but described structural fragmentation hindering its delivery, including incompatible IT systems, unclear roles and weak communication pathways. These gaps left non-clinical staff underused, GPs overstretched, and people with dementia and carers unsupported by a joined-up approach.

Our findings align with previous research positioning personalised dementia care as relational and shaped by communicative competence and organisational context [14]. The cross-cutting themes of time and conversation as intervention highlight that communication is not an adjunct but the foundation of care planning. Yet the organisational conditions required to sustain this work remain inconsistent and fragile.

Evidence on ARRS role integration highlights uncertainty around remit, communication pathways, and access to patient information [26,27]. We extend this by showing the communicative consequences of such ambiguity, including reduced continuity, fewer opportunities for shared understanding, and limited co-creation of meaningful care plans.

While previous work notes reliance on carers to facilitate communication [11], our findings show that carers often experience this as a burden, particularly when they feel responsible for interpreting needs or when inaccessible formats such as phone calls are used. This aligns with earlier research showing that language barriers, including reliance on telephone interpreters, can hinder rapport and the communication of empathy [28].

Across all groups, participants were also uncertain about what a ‘care plan’ is, whether one existed, and who was responsible for it, reinforcing evidence that care plans are often invisible in practice [8]. Greater conceptual clarity is therefore foundational to meaningful personalised planning and shared ownership.

### Implications for practice

Policy frameworks [29,30] prioritise personalised conversations, shared decision-making, and adaptable care plans to prevent unnecessary admissions. However, our findings suggest the communicative foundations required to realise these ambitions remain weak.

NHS *Fit for the Future* [5] emphasises digital upskilling. While this may reduce administrative burden, it risks sidelining workforce training in communicative competencies. Despite guidance from the Personalised Care Institute [12] and the Dementia Training Standards Framework [31], practitioners reported uncertainty about adapting communication and managing triadic interactions. Digital transformation should be accompanied by investment in conversational skills.

Cultural competence must extend beyond awareness to include communicative strategies that preserve dignity and sustain participation. Clearer shared understanding of care plan purpose and ownership is also needed. Co-designed, accessible preparatory tools could support this.

Crucially, effective use of the expanded workforce requires capitalising on untapped potential. Non-clinical roles were widely perceived as having the time and relational orientation required for personalised dementia care yet remained structurally underused. Their integration into multidisciplinary care planning is therefore essential.

### Policy implications

Policy ambitions for personalised dementia care depend on communicative practices that remain under-resourced. While workforce expansion has been prioritised, integration has lagged behind. Policies should move beyond role creation to formal inclusion, giving non-clinical staff the authority, access, and training to lead care planning. Without this, the relational capacity of the workforce will continue to be underused.

### Strengths and limitations

A principal strength of this study is its multi-perspective design, capturing how care planning conversations are experienced by people with dementia, informal carers, and a diverse range of practitioners. By foregrounding communicative processes in primary care and earlier-stage dementia, the study addresses key evidence gaps and advances understanding of the relational foundations of personalised care. A limitation is the underrepresentation of marginalised groups, particularly people from minority ethnic backgrounds and diverse sexual orientations, thereby constraining transferability. Future research should prioritise underserved populations, use observational methods to examine real-time care planning, and evaluate system-level innovations such as shared digital care plans to support communicative continuity.

## Conclusion

This study highlights a persistent gap between policy ambitions for personalised dementia care and everyday practice. Time-poor, checklist-driven encounters and fragmented systems undermine the trust-building conversations that enable disclosure and shared decision-making. Closing this gap requires investment in communicative skills alongside structural reform, particularly through integrating the wider primary care workforce and its underused relational capacity.

## Data Availability

All data produced in the present study are available upon reasonable request to the authors. Due to the sensitive nature of the qualitative data collected for this study, and the
ethical restrictions associated with participant confidentiality, the datasets generated
and analysed are not publicly available. De-identified interview transcripts may be
made available to bona fide researchers for secondary analysis, subject to appropriate
ethical approvals and assessment of suitability. Requests for access should be
directed to the corresponding author at.
Access will be considered by the authorship team in line with the original ethical
approvals and data governance arrangements.

